# ACE2 Polymorphisms rs2285666 and rs147311723 Are Clinically Relevant Variants in SARS-CoV-2 and *Mycobacterium tuberculosis* Co-infection

**DOI:** 10.64898/2026.09.11.26362837

**Authors:** Mary Ngongang Kameni, Assam Assam Jean Paul, Eric Berenger Tchoupe, Noubissi Toyim Albert, Ange Maxime Tchoutang, Maloba Franklin, Taya Fokou Jean Bosco, Tsague Voufo Svetlana Carel, Georgette Njila, Fuh Roger Neba, Severin Donald Kamdem, Palmer Masumbe Netongo

## Abstract

**Background:** Severe acute respiratory syndrome coronavirus 2 (SARS-CoV-2) enters host cells primarily through the angiotensin-converting enzyme 2 (ACE2) receptor. Genetic variation in the ACE2 gene may influence host inflammatory responses and clinical outcomes during COVID-19. Tuberculosis (TB) on the other hand increases the level of ACE2 expression through interferon stimulation. The objective of this study was to investigate the association between specific single-nucleotide polymorphism (SNP) in the ACE2 gene and their impact on susceptibility to and severity of SARS-CoV-2 and *Mycobacterium tuberculosis* Co-infection.

**Methods:** This retrospective cross-sectional study included 120 symptomatic individuals recruited between September 2020 and December 2023 from four hospitals in Yaoundé, Cameroon. Participants were classified into four groups: COVID-19, TB, TB/COVID-19 co-infection, and infection-negative controls. Diagnoses were confirmed using RT-PCR and standard microbiological methods. Serum biochemical markers and cytokines were quantified, and genotyping of ACE2 polymorphisms was performed using TaqMan assays.

**Results:** Statistical analysis showed a significant association between ACE2 expression and inflammatory or biochemical markers, particularly for rs147311723. Several ACE2 polymorphisms (rs6632677, rs147311723, rs4646140, rs2285666, and rs4646142) were significantly associated with increased pro-inflammatory cytokine levels. Selected biochemical parameters demonstrated suggestive associations with ACE2 variants, including creatinine (rs147311723, p = 0.0522), AST (rs2285666, p = 0.0536), and ALT (rs4646142, p = 0.0582), indicating potential renal and hepatic involvement. Principal component analysis identified distinct immunometabolic profiles across patient groups, with co-infected individuals exhibiting heightened inflammatory and metabolic perturbations.

**Conclusions:** These findings suggest that ACE2 genetic variability is associated with modulation of inflammatory responses and may contribute to organ-specific dysfunction in COVID-19 and TB across different populations. This highlights the potential role of host genetics in shaping disease severity and underscore the need for larger studies to validate these associations in diverse populations.

## Introduction

Severe acute respiratory syndrome coronavirus 2 (SARS-CoV-2), the virus that causes coronavirus disease 2019 (COVID-19) via respiratory infection, was first discovered in Wuhan, China, in December 2019 (Wang *et al*., 2020). The SARS-CoV-2 virus was first identified in Huanan, China, and has caused a tremendous number of cases and deaths worldwide. (Hou *et al*., 2020; Shapira *et al*., 2022). Numerous studies have identified several risk factors linked to COVID-19 susceptibility and /or severity. These include gender, older age, specific ethnicities, co-infections, and respiratory disorders. Additionally, host genetic factors have demonstrated a significant influence on susceptibility to infection and disease severity (Horowitz *et al*., 2022) . Mycobacterium tuberculosis (Mtb), on the other hand, is the bacterium responsible for tuberculosis (Kameni *et al*., 2025). The modes of transmission, symptoms, and manifestation of both SARS-CoV-2 and Mtb are quite similar, and their mechanism of infection have been linked to two principal proteins in the body: the Angiotensin Converting Enzyme 2 (ACE-2) and the Transmembrane Protease Serine 2 (TMPRSS2) (Abdelsattar *et al*., 2022). Human host cells’ invasion by SARS-CoV-2 depends on the presence of these two key factors. ACE2 serves as the receptor through which the virus enters the host cells, while TMPRSS2 facilitates the priming and activation of the viral spike protein, enabling viral fusion and entry (Jackson *et al*., 2022). ACE2 helps regulate blood pressure and features a single transmembrane domain and is mainly expressed in the heart, kidney, and lungs (Burrell *et al*., 2004) . Whilst TMPRSS2 is a type II transmembrane serine protease with a catalytic triad (His296, Asp345, Ser441) involved in cleaving substrates for physiological processes (KOYOU et al., 2025; Qu et al., 2024; Sonawane et al., 2021). It is expressed on the surface of respiratory epithelial cells. Due to the significance of ACE2 in the viral entry process, it is important to understand the genetic variations in ACE2 and their association with COVID-19 outcomes. A multitude of conflicting studies explored the effects and prevalence of genetic variations within the ACE2 gene and their association with COVID-19. Some studies found that an increase in ACE2 level is correlated with severe cases of COVID-19, in conjunction with specific laboratory parameters (Abdelsattar *et al*., 2022; Zheng, 2022). This could be linked to altered immune response, which may activate pro-inflammatory pathways, thus exacerbating lung injury (Xiao *et al*., 2020). In Tuberculosis, Mtb infection upregulates ACE2 expression in lung tissues via interferon stimulation (Shah *et al*., 2022). This mechanism is also essential for SARS-CoV-2 entry and replication. Single-nucleotide polymorphisms (SNPs) significantly affect the interaction between microbial agents and host cells, the body’s resistance to infection, and the manifestation of severe disease symptoms (Meseldžić *et al*., 2024). Numerous studies have revealed that the frequencies of certain alleles and SNPs in these genes are associated with differences in COVID-19 and TB prevalence and susceptibility among various ethnic groups (Srivastava *et al*., 2020; Abdelsattar *et al*., 2022; Li *et al*., 2022; Zheng, 2022; Kameni *et al*., 2025). Understanding the role of this protein is crucial for developing treatments and interventions to mitigate the spread of the virus and its impact on the human body. Consequently, the aim of the present study is to investigate the roles of Ace2 rs4646179 A>G, rs147311723 G>A, rs4646142 G>A, rs2074192 C>T, rs35803318 C>T, rs4646140 C>T, rs6632677 G>C, rs4646116 T>C, rs2285666 C>A, rs4240157 C>G in the susceptibility and severity of COVID-19 and TB co-infection concerning clinical findings and laboratory investigations in a Cameroonian population.

## Materials and Methods

### Study Population

All study participants recruited presented signs and symptoms of a respiratory disease. Every participant underwent clinical examination and laboratory assessment for COVID-19, TB, and other infectious diseases endemic in the region, such as Influenza A and B, malaria, HIV, and Hepatitis B. A group of symptomatic individuals who tested negative for COVID-19, TB, and the other infectious diseases mentioned above, who had been screened during the study, were enrolled as controls. Patients who tested positive for TB and/ or COVID-19 (confirmed by real-time polymerase chain reaction [RT-PCR] test), as well as a control group of COVID-19 and TB negative patients, were included in this study.

### Study Design and Subject Criteria

This retrospective cross-sectional study comprised a total of 120 participants. Between September 2020 to December 2023, participants were recruited from four distinct hospital facilities in Yaounde, Cameroon. Djoungolo Hospital, Ekoumdoum Baptist Hospital, Red Cross Hospital, and Yaoundé Jamot Hospital. The study participants were classified into four groups: COVID-19 positive, TB positive, TB-COVID-19 association, and controls (Table 1).

### Data collection

Blood specimens were collected by standard venipuncture. After coagulation at room temperature for 30 min, serum was separated by centrifugation at 5000 g for 10 min and stored at -20 °C until analysis. Serum samples were aliquoted and stored to ensure their availability for specialized analyses conducted as part of this project. Plasma was obtained from blood collected in EDTA tubes, aliquoted, and stored at - 80 °C for future use.

Sputum samples were collected using plastic cups with a 40 mL capacity. After collection, sputum microscopy using the Ziehl-Neelsen staining technique was carried out. A confirmatory real-time PCR using the SARAGENE™ Mycobacterium tuberculosis test, COSARA Diagnostics Ltd, India, and LogixSmart MtbKit (Cat #: MTB-K-007)-Co-Diagnostics Inc, USA was performed on extracted bacterial DNA according to the manufacturer’s instructions.

Nasopharyngeal samples were collected from the participants by inserting the swab provided about 2- 2.5 cm into the nostrils. The HIGHTOP Antigen Rapid Test device that Qingdao Hightop Biotech Company manufactured was used according to the manufacturer’s instructions. The QIAamp viral RNA mini kit extracted coronavirus RNA from nasopharyngeal samples. A confirmatory COVID-19 diagnosis was later made using real-time PCR using the Logix Smart ABC (Cat #: ABC-K-001) test utilizing the patented Co-Primer technology according to the manufacturer’s instructions. The Co-Primer triplex assay uses extracted viral RNA to detect Influenza A, B, and SARS-CoV-2 (gene RdRp and E-gene) in upper respiratory tract samples, including saliva (Netongo et al., unpublished data).

### Biochemical and hematological measurements

Liver and kidney function markers were assessed in serum samples using spectrophotometric methods. Levels of AST, ALT, bilirubin (direct and total), urea, and creatinine were measured using reagent kits from Precise Max reagent kit according to the manufacturer’s instructions and Genuine Biosystem (WWHS Biotech. IncShenzhen, P.RChina). Three analytical approaches were applied: the kinetic method, which monitored changes in absorbance over time to quantify creatinine, AST, and ALT; the endpoint method, which measured bilirubin concentration upon the completion of the reaction; and the Berthelot method, which quantified urea by converting ammonia produced from urea hydrolysis into a colored indophenol compound through reaction with phenol and hypochlorite ions under alkaline conditions.

### Measurement of inflammatory cytokines and Circulating ACE2 level

Serum cytokines (IL-6, IFN-γ, TNF-α, IL-10, IL-2, and IL-1β) were assayed in serum using sandwich ELISA Origene kits (Origene Technologies, Inc, Rockville, MD20850, US) according to the manufacturer’s instructions. ACE2 levels were measured in plasma samples using specific ELISA kits from RayBiotech (ACE2 Cat#: ELH-ACE2) (Lui *et al*., 2020).

### Genotyping to detect mutation in SNPs of ACE2 gene

Genomic DNA was harvested from the peripheral blood using the commercially available Quick-DNA™ Miniprep Kit (QIAamp DNA Blood Mini kit, Qiagen, Germany), according to the manufacturer’s instructions, and its quality was verified in agarose gels stained with ethidium bromide nucleic acid gel stain (Thermo Fisher Scientific, C.A, USA). Then, the DNA concentration was measured using a nanodrop (Thermo Fisher Scientific, C.A, USA), and purity was determined by calculating the A260/280 ratio. The extracted genomic DNA was stored at -80°C until used in the genotyping reaction. SNPs were analyzed by real-time polymerase chain allelic discrimination technology using TaqMan SNP genotyping assay kit (Thermo Fisher Scientific, Waltham, MA, United States) on a Co-Dx Box Magnetic Induction Cycler qualitative Time Polymerase Chain Reaction (qPCR) machine (Co-Diagnostics Inc, USA, Cat # MIC001355). Ten variants were analyzed for the Angiotensin converting enzyme gene namely, rs2285666, rs4240157, rs4646142, rs4646116, rs6632677, rs4646140, rs147311723, rs2074192, rs35803318, and rs4646179. Specifically, genotype variants were determined using the TaqMan™ SNP Genotyping Master Mix kit from Thermo Fisher Scientific, C.A, USA (Cat #: 4381656) that reveals Ace2 rs4646179 A>G, rs147311723 G>A, rs4646142 G>A, rs2074192 C>T, rs35803318 C>T, rs4646140 C>T, rs6632677 G>C, rs4646116 T>C, rs2285666 C>A, rs4240157 C>G (Table 2). The reaction mix of each sample was composed of 5 µL of 2X TaqMan Genotyping Master Mix, 0.5 µL of TaqMan assay (20X), and 4.5 µL RNase-free water. The thermal cycling protocol is optimized at 95°C for 10 min for AmpliTaq Gold, UP Enzyme Activation, followed by a denaturation step at 95°C for 15 s and annealing/extension at 60°C for 1 min for 40 cycles. The qPCR was performed on a Co-Diagnostics PCR instrument (Co-Diagnostic, INC, Salt Lake City, USA), and the results were analyzed using Co-Diagnostic genotyper software. This software was used to plot the findings of the allelic discrimination data as a scatter plot of Allele 1 (VIC® dye) versus Allele 2 (FAM™ dye). Each well of the 96-well reaction plate was represented as an individual point on the plot.

### Statistical analysis

The statistical analysis followed a structured and integrative approach. Preliminary tests were first applied to assess the normality of distributions (Shapiro-Wilk), homogeneity of variances (Levene’s test), and absence of collinearity between variables. Associations between ACE2 SNP genotypes and clinical groups were then examined using Fisher’s exact test. Comparisons of immunological and biochemical markers by genotype were performed using the non-parametric Kruskal-Wallis test. Pearson correlation was used to explore linear relationships between parameters. Finally, a principal component analysis (PCA), including biplot and cos², was used to examine the overall structure of the data. All analyses were performed using R version 4.5.2 with specialized packages.

## Results

### Demographic and clinical features of study participants

Every study participant underwent clinical examination and laboratory assessment for COVID-19 and TB. In total, only 119 participants were included in this study. The study populations consisted of four groups: COVID-19 positive (n= 31), TB positive (n= 43), TB COVID-19 positive (n= 21) and a set of controls (n= 24). The control group consisted of participants who tested negative for all the above-mentioned diseases screened at the time of the study. The proportion of males to females was 60% and 40%, respectively. The most frequent symptoms in the COVID-19 group (n=31) were cough (74%), fever (70.9%) and headache (61.3%). The main clinical signs reported during TB-COVID-19 association (n=21) were cough (85.7%), fatigue (80.9%) and headache (76.1%). TB patients (n=43) presented predominant symptoms such as cough (53.5%), fever (46.6%) and chest pain (25.6%). Details of sociodemographic and clinical data are summarized in Table 1.

Significant association between rs6632677, rs147311723, rs4646140, rs2285666, and rs4646142 of the ACE2 gene and pro-inflammatory markers expression levels.

Kruskal-Wallis analysis showed that ACE2 polymorphisms primarily influence pro-inflammatory cytokine production. TNF-α showed the strongest and most consistent associations, being linked to three independent SNPs: rs6632677 (p = 0.0091), rs2285666 (p = 0.0194), and rs4646142 (p = 0.0472). IL-6, another key inflammatory mediator, was associated with rs147311723 (p = 0.0471) and rs4646140 (p = 0.0364). IFN-γ, on the other hand, was associated with rs2285666 (p = 0.0465). IL-1β, linked to rs6632677 (p = 0.0454), further supports the role of ACE2 variants in inflammasome activation and pyroptotic responses (Figure 1).

**Figure 1:**
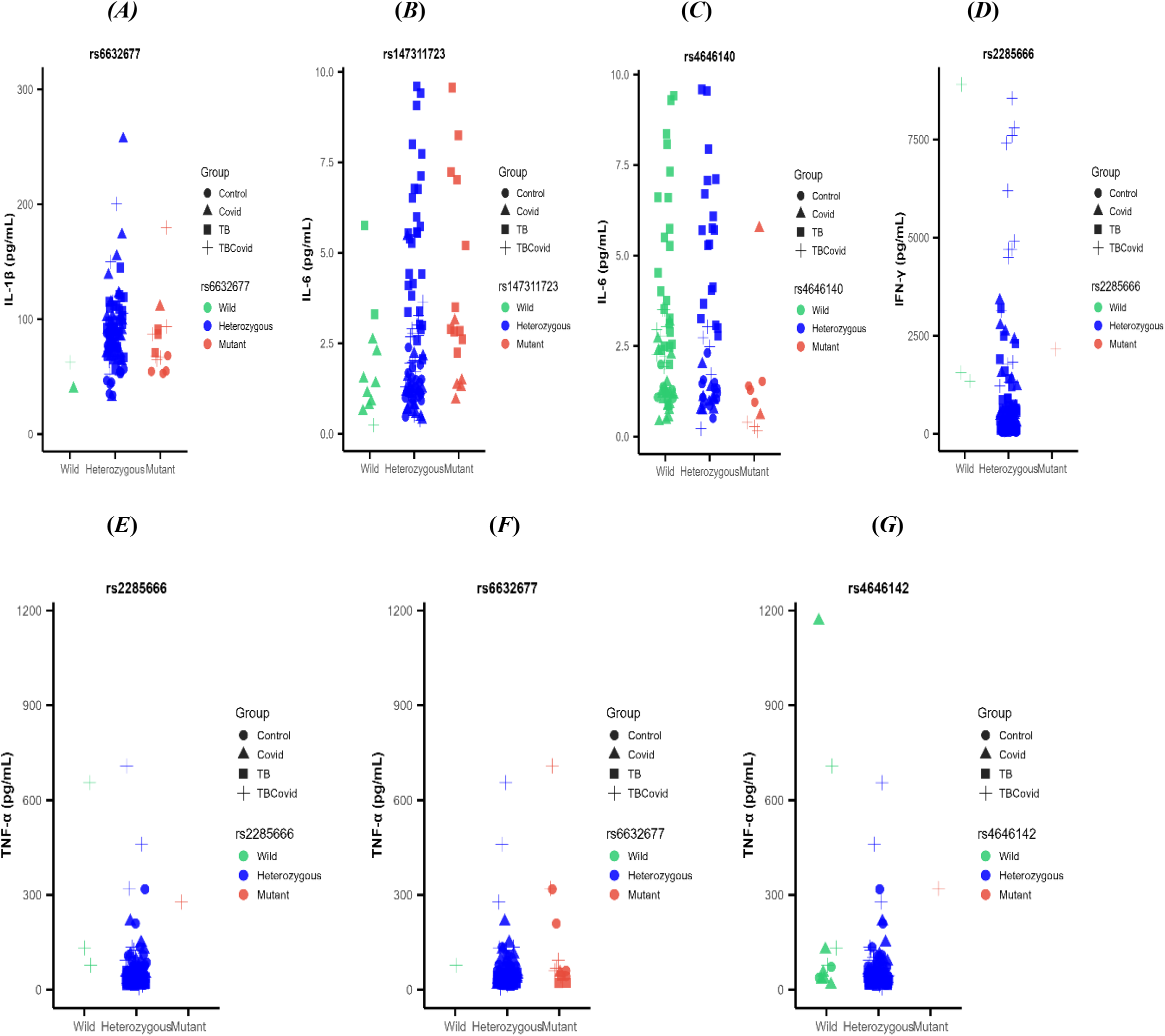
Kruskal–Walli’s analysis used for comparing three or more categorical groups revealed a significant association between five ACE2 gene SNPs (rs66223677, rs147311723, rs4646140, rs2285666, and rs4646142) and the expression levels of pro-inflammatory cytokines (IL-1β, IL-6, IFN-γ, TNF-α). The x-axis indicates the genetic profile while the y-axis measures cytokine levels. Each data point represents an individual, with shapes indicating group membership: circles (●) = negative control group, triangles (▴) = COVID-19⁺ group, squares (▪) = tuberculosis (TB)⁺ group, and plus signs (+) = TB and COVID-19 co-infection. Colours denote mutation type: green = wild-type allele, blue = heterozygous mutant allele, red = double mutant allele.

Significant association between rs2285666, rs4646142, and rs147311723 of the ACE2 gene and some biochemical marker levels.

Three biochemical parameters (Creatinine; rs147311723, p = 0.0522, AST; rs2285666, p = 0.0536 and ALT; rs4646142, p = 0.0582) showed statistically significant associations with SNPs of ACE2. This suggests potential renal vulnerability, consistent with the role of ACE2 in renal physiology and susceptibility to acute kidney injury during severe infections. It also indicates possible hepatic involvement through inflammation-mediated tissue injury (Figure 2).

**Figure 2:**
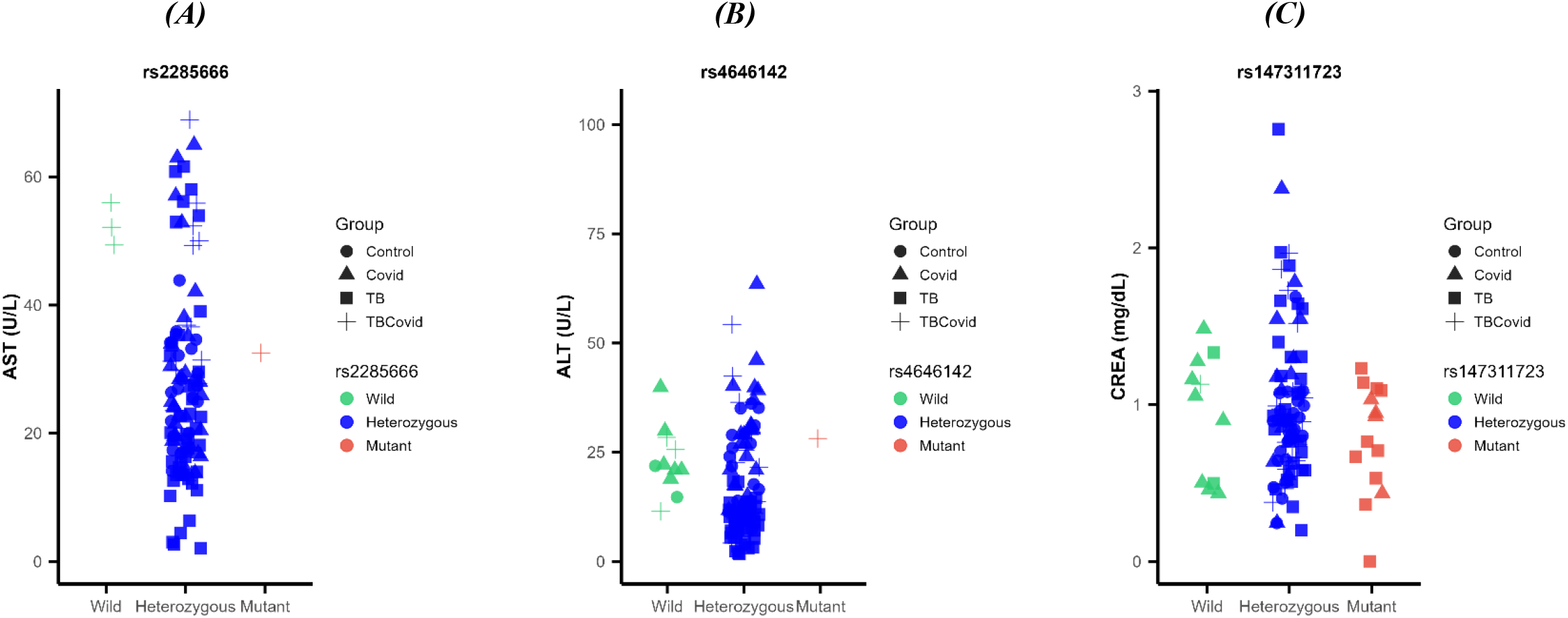
Kruskal–Walli’s analysis used for comparing three or more categorical groups revealed a significant association between three ACE2 gene SNPs (rs2285666, rs4646142 and rs147311723) and the expression levels of biochemical markers (AST, ALT, CREA). The x-axis indicates the genetic profile while the y-axis measures biochemical marker levels. Each data point represents an individual, with shapes indicating group membership: circles (●) = negative control group, triangles (▴) = COVID-19⁺ group, squares (▪) = tuberculosis (TB)⁺ group, and plus signs (+) = TB and COVID-19 co-infection. Colours denote mutation type: green = wild-type allele, blue = heterozygous mutant allele, red = double mutant allele.

Weak positive correlation between creatinine, IL1-β, total bilirubin, and ACE2 levels in rs147311723 polymorphism of ACE2.

The Rs147311723 polymorphism of ACE2 emerged as one of the most clinically relevant ACE2 variants. The Pearson correlation analysis was performed between biochemical markers and cytokines for participants with COVID-19, TB, and TB/COVID-19. A weak positive correlation was observed between creatinine and ACE2 expression (r = 0.321, p = 0.0359) in TB-positive patients, as well as between IL-1β and ACE2 levels (r = 0.45, p = 0.0111) in COVID-19 patients. On the other hand, a weak negative correlation (r = -0.5, p = 0.0192) was observed between total bilirubin and ACE2 expression levels in TB/COVID-19 co-infected patients (Figure 3).

**Figure 3:**
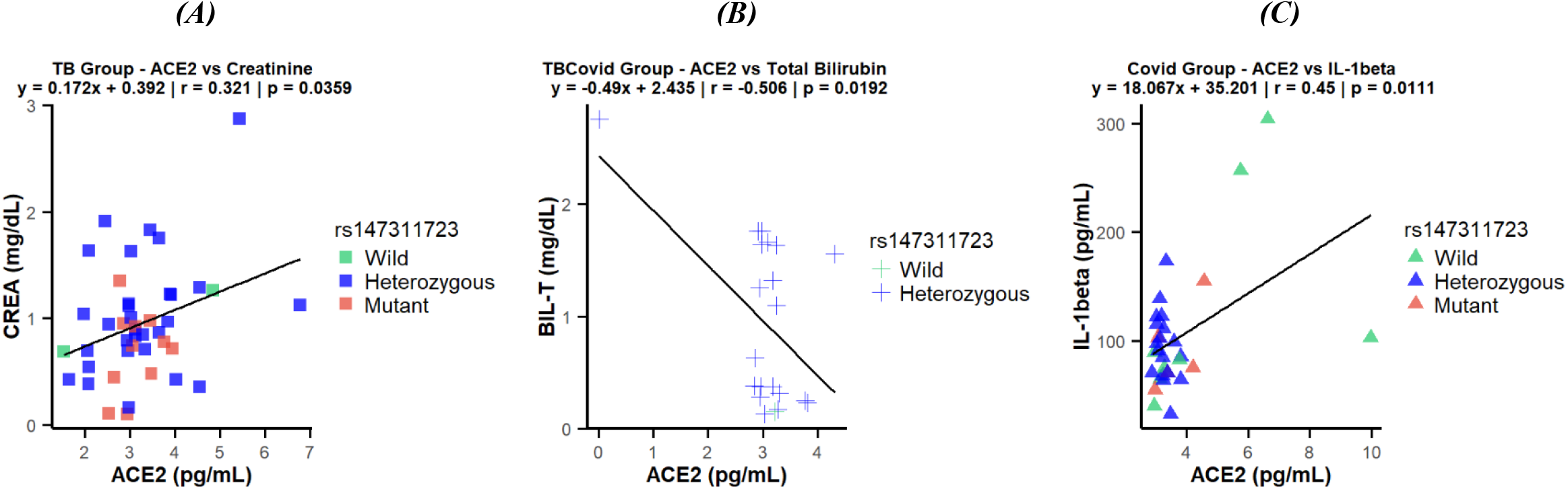
Pearson correlation analysis showed that interactions are stronger when data points lie closer to the linear regression line. The following weak correlations were observed for the rs147311723 polymorphism of the ACE2 gene: A) Weak positive correlation between creatinine and ACE2 levels in TB + patients. B) Weak negative correlation between total bilirubin and ACE2 expression levels in TB and COVID-19 co-infected patients. C) Weak positive correlation between IL-1β and ACE2 levels in COVID-19+ patients.

Correlation between biochemical markers and cytokine expression across the study population.

Correlations between biochemical markers and cytokine expression were assessed across the study population. Urea, IFN-γ, TNF-α, AST, and ALT exhibited positive correlations, with a particularly strong association observed between IFN-γ and TNF-α (p = 0.00044). Similarly, IL-6 and D-dimer showed a positive correlation (p = 0.01), while ACE2, IL-4, IL-10, and IL-1β were also positively correlated (p < 0.05), with a particularly strong correlation observed between IL-2 and IL-1β. However, IL-4 demonstrated weaker interactions compared to the other interleukins. In contrast, D-dimer demonstrated negative correlations with urea, IFN-γ, TNF-α, AST, and ALT. No significant correlation was found between IL-6 and ALT. The correlation circle (Cos² plot) graph further supported these findings, with positively correlated variables appearing closer to each other on the Cos² graph (Figure 4).

**Figure 4:**
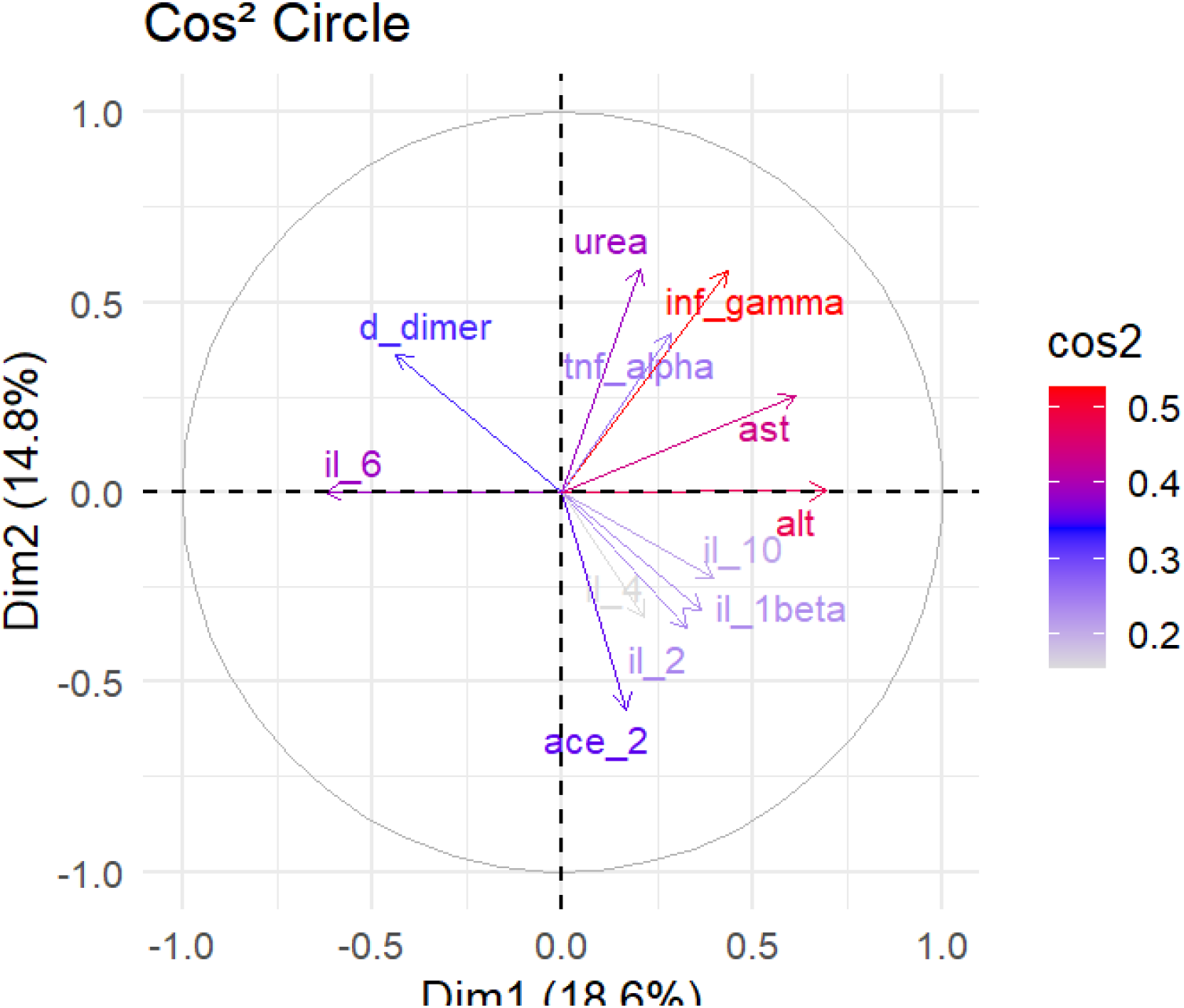
Correlation circle (Cos² plot) shows the multidimensional relationships between various clinical and inflammatory biomarkers. Positively correlated variables appear closer to each other and point in the same direction on the Cos² plot. The color scale ranges numerically from 0.2 to 0.5 to indicate the strength of this representation. Red vectors denote the most influential variables while arrows tending towards blue or pale gray have a weak contribution on this plane.

Principal component analysis reveals significant correlations between biochemical markers and cytokine expression between TB, COVID-19, and TB/COVID-19 co-infected patients.

A principal component analysis (PCA) biplot was generated to visualize sample clustering, the contributions of variables to the principal components (PCs), and correlations between biomarkers. In this representation, individual samples are plotted as points in the reduced PCA space, while vectors (arrows) represent the direction and strength of the contribution of each original variable to the PCs. The X- and Y-axis correspond to the first two principal components, which explained 33.4% of the total variance. Key drivers of variation along PC1 included ALT, IL-6, AST, and D-dimer, while urea, IFN-γ, ACE2, and TNF-α were the most influential variables on PC2. Stratified by patient groups, TB positive patients were characterised by elevated levels of IL-6 (in a predominant subgroup) and D-dimer (in a smaller subgroup). TBCOV patients showed increased concentrations of IFN-γ, AST, urea, and TNF-α, along with moderate elevation in IL-10 (in a smaller subgroup). COVID-19 patients displayed a distinct immunometabolic profile, marked by significantly higher levels of urea, TNF-α, ALT (in a smaller subgroup), ACE2, IL-2, IL1-β and IL-10 (in a predominant subgroup) compared to the other groups. The control group had no significant effect on the variation of biochemical markers or interleukin levels. In contrast, IL-4 levels remained stable across all groups, including controls, TB, COV, and TBCOV (Figure 5).

**Figure 5:**
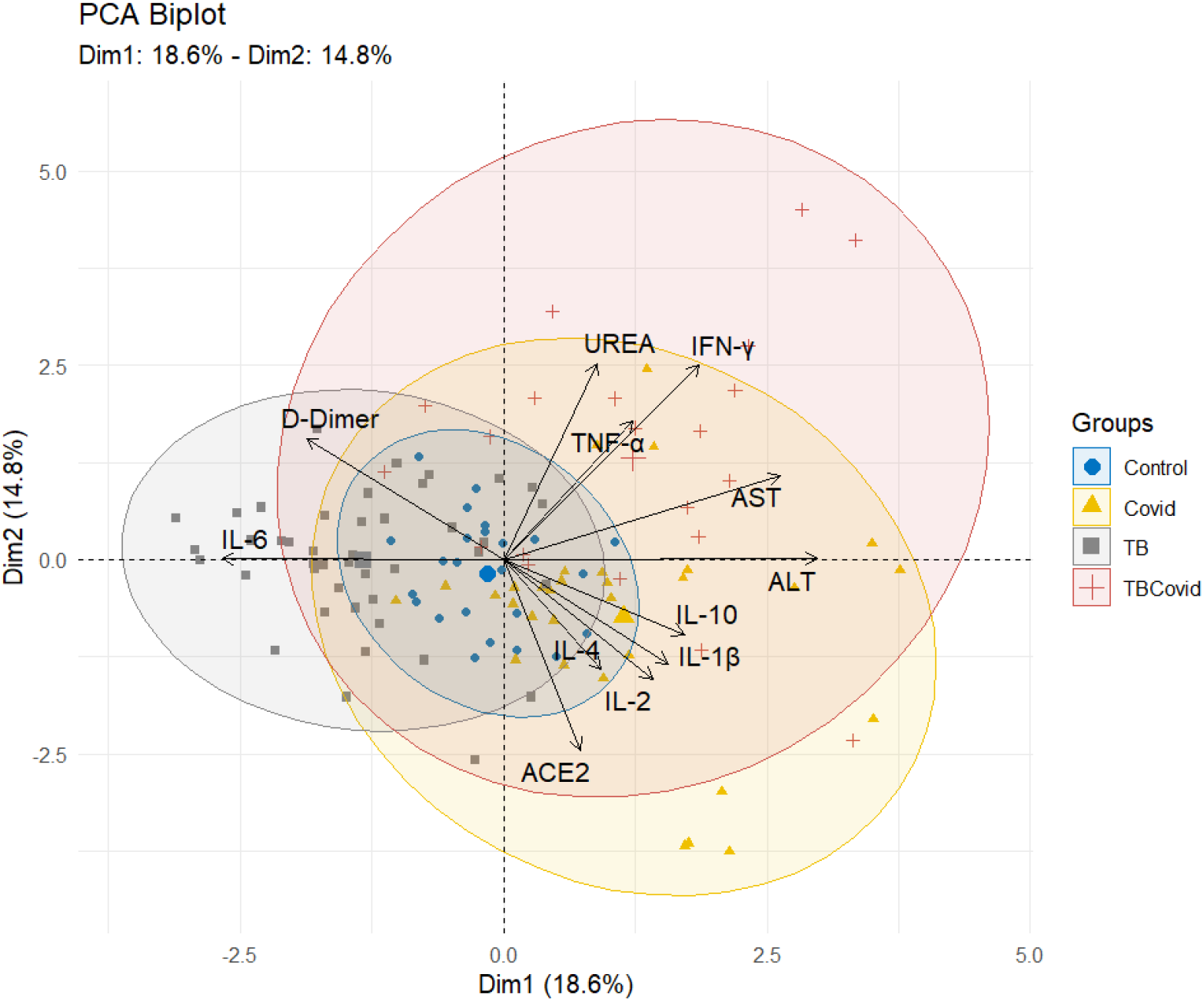
The Principal Component Analysis (PCA) biplot, with Dim1 and Dim2 accounting for 18.6% and 14.8% of the total variance respectively, reveals the segregation of clinical profiles based on biomarker expression. The confidence ellipses clearly discriminate the cohorts: the control group (blue) clusters at the origin (baseline profile); the tuberculosis group (grey) segregates negatively along Dim1, associated with IL-6 and D-Dimers; the COVID-19 group (yellow) projects into the lower right quadrant, correlated with transaminases (AST, ALT) and interleukins (IL-1β, IL-2, IL-10). Finally, the TBCovid co-infection (red) exhibits the highest dispersion (upper right quadrant), highlighting a severe and heterogeneous hyper-inflammatory profile dominated by IFN-γ, TNF-α, and UREA.

## Discussion

ACE2 polymorphisms, notably rs147311723 and rs2285666, are strongly associated with susceptibility to COVID-19, tuberculosis, and TB-COVID-19 co-infection. The absence of the mutant rs147311723 genotype in co-infected individuals suggests a protective role, while the exclusive presence of the wild-type rs2285666 homozygote in this group may be a critical determinant of outcome (Chukkayapalli *et al*., 2023). Other SNPs such as rs4646142, rs4646140, and rs6632677 showed moderate associations, highlighting a complex genetic influence (Hou *et al*., 2020). In contrast, rs2074192 shows no association, confirming the specificity of these genetic effects on pathogenesis. ACE2 gene polymorphisms significantly influence pro-inflammatory cytokine production. Kruskal-Wallis analyses showed that TNF-α has the strongest and most consistent associations with multiple polymorphisms, including rs6632677 (p = 0.0091), rs2285666 (p = 0.0194), and rs4646142 (p = 0.0472). This convergence strongly implicates ACE2 genetic variation in regulating TNF-α-mediated inflammation, a major component of the cytokine storm observed in diseases such as COVID-19 and tuberculosis **(Badawi, 2020; Admou, 2021)**. Interleukin-6 (IL-6), another essential pro-inflammatory cytokine, is associated with rs147311723 (p = 0.0471) and rs4646140 (p = 0.0364) polymorphisms. Given the role of IL-6 in acute phase responses and disease severity (Deng *et al*., 2024; Feira *et al*., 2025) these associations suggest that ACE2 variants may influence clinical outcomes by modulating inflammatory cascades (Costela-Ruiz *et al*., 2020). The association of IFN-γ with rs2285666 (p = 0.0465) is particularly relevant for anti-tuberculosis immunity, as this Th1 cytokine induces macrophage activation and granuloma formation (Muller *et al*., 2013). IFN-γ is also involved in the antiviral response and may contribute to severe pulmonary inflammation in COVID-19 (Costela-Ruiz *et al*., 2020). Moreover, IL-1β, linked to rs6632677 (p = 0.0454), is a potent mediator of inflammation and fever. Regarding biochemical markers, three parameters showed associations close to significance. Creatinine (rs147311723, p = 0.0522) suggests potential renal vulnerability, consistent with the role of ACE2 in renal physiology and susceptibility to acute kidney injury during severe infections (Vergara *et al*., 2022). AST (rs2285666, p = 0.0536) and ALT (rs4646142, p = 0.0582) indicate possible hepatic involvement through inflammation-mediated tissue injury (Cai *et al*., 2020). Studies on ACE2 polymorphisms have also demonstrated their association with COVID-19 severity, particularly in men (Martínez-Gómez *et al*., 2022). Genetic variants in ACE2 and TMPRSS2 genes are linked to susceptibility and severity of COVID-19 infection (Makled *et al*., 2023; Meseldžić *et al*., 2024). The rs147311723 polymorphism of the ACE2 gene is clinically relevant due to its positive correlations with biochemical markers (creatinine in tuberculosis patients) and immunological markers (IL-1β in COVID-19 patients). Conversely, a weak negative correlation is observed between total bilirubin and ACE2 expression in patients co-infected with tuberculosis and COVID-19 (Martínez-Gómez *et al*., 2022) . These findings underscore the influence of ACE2 variants on inflammatory response and organ function in the context of COVID-19 and tuberculosis (Badawi, 2020; Admou, 2021). Correlation (cos²) and principal component analyses revealed distinct immunometabolic signatures characterizing COVID-19, tuberculosis, and co-infection. The strong positive correlation between IFN-γ and TNF-α (p = 0.00044) reflects coordinated Th1-type inflammatory responses, central to both antiviral and antimycobacterial immunity (Cooper and Mayer-Barber, 2011; Gardinassi *et al*., 2020). These cytokines, combined with elevated urea and transaminase levels, suggest a common pathophysiology involving systemic inflammation and tissue injury (Chen *et al*., 2020; Pedersen and Ho, 2020). The positive correlation between IL-6 and D-dimer (p = 0.01) indicates hyperinflammation, characteristic of severe forms of COVID-19 (Connors and Levy, 2020; Satre Buisson, 2020). D-dimer is an essential biomarker of COVID-19-associated coagulopathy (CAC), characterized by elevated D-dimer and fibrinogen levels, requiring an anticoagulation strategy adapted to disease severity. Stratification by PCA reveals specific profiles. Tuberculosis patients show an IL-6/D-dimer predominance, reflecting chronic inflammation and increased thrombotic risk (Duffy *et al*., 2019). TB/COVID-19 co-infection is characterized by elevations in IFN-γ, TNF-α, and transaminases, indicating severe systemic inflammation with organ involvement (Costela-Ruiz *et al*., 2020; Xiao *et al*., 2020). COVID-19 with elevated ACE2, IL-2, IL-1β, IL-10, and urea levels demonstrates disease severity associated with multi-organ failure (Lucas *et al*., 2020; Rahman *et al*., 2020; Huang *et al*., 2022). The stability of IL-4 across all groups indicates the maintenance of Th2 responses despite Th1/inflammatory response dominance (Grifoni *et al*., 2020; Hajjo and Tropsha, 2020). These distinct signatures underscore the importance of personalized therapeutic targeting based on immunometabolic phenotyping.

## Conclusion

The convergence of genetic disease associations (p < 0.001) and correlations with immunological biomarkers (p < 0.05) provides cross-validated evidence of genuine biological effects. Rs2285666 and rs147311723 emerge as the most clinically relevant variants, influencing both disease susceptibility and inflammatory responses. The unique genotypic profiles observed in TB/COVID co-infection suggest that these polymorphisms could guide risk stratification for dual infections. These findings support potential therapeutic implications, including personalized targeting of the renin-angiotensin system or ACE2 genotype-based anti-inflammatory interventions, although further validation in larger cohorts remains necessary.

## Supporting information

https://docs.google.com/document/d/1Qg9nVQFBlbtU1rxPrbDtTWPhalmwwtRB/edit?usp=sharing&ouid=108297540105191031878&rtpof=true&sd=true

## Ethics statement

This study, which included human participants, complied with the ethical standards established in the 1964 Helsinki Declaration and received approval from the Cameroon National Ethical Committee for Research in Human Health

(N° 2020/07/1265/CE/CNERSH/SP) in Yaoundé. Written informed consent was obtained from all participants, with confidentiality assured and measures taken to minimize risks and adverse effects.

## Data Availability Statement

Data will be made available on request

## Author’s Contributions

**Conceptualization:** Palmer Masumbe Netongo, Mary Ngongang Kameni, Eric Berenger Tchoupe, Severin Donald Kamdem

**Funding acquisition:** Palmer Masumbe Netongo

**Methodology:** Mary Ngongang Kameni, Eric Berenger Tchoupe, Severin Donald Kamdem

**Investigation:** Mary Ngongang Kameni, Eric Berenger Tchoupe, Severin Donald Kamdem

**Software:** Noubissi Toyim Albert, Mary Ngongang Kameni, Georgette Njila

**Data Curation:** Mary Ngongang Kameni, Eric Berenger Tchoupe, Severin Donald Kamdem

**Project Administration:** Palmer Masumbe Netongo, Assam Assam Jean Paul

**Laboratory analysis:** Mary Ngongang Kameni, Eric Berenger Tchoupe, Fuh Roger Neba

**Statistical analysis:** Noubissi Toyim Albert, Ange Maxime Tchoutang, Mary Ngongang Kameni, Georgette Njila

**Supervision:** Palmer Masumbe Netongo, Assam Assam Jean Paul

**Writing – original draft:** Mary Ngongang Kameni, Eric Berenger Tchoupe, Ange Maxime Tchoutang, Noubissi Toyim Albert, Tsague Voufo Svetlana Carel, Franklin Maloba, Georgette Njila, Palmer Masumbe Netongo

**Writing – review & editing:** Mary Ngongang Kameni, Assam Assam Jean Paul, Eric Berenger Tchoupe, Noubissi Toyim Albert, Ange Maxime Tchoutang, Maloba Franklin, Taya Fokou Jean Bosco, Tsague Voufo Svetlana Carel, Georgette Njila, Fuh Roger Neba, Severin Donald Kamdem, Palmer Masumbe Netongo

