## Supplementary material for "ACE2 Polymorphisms rs2285666 and rs147311723 Are Clinically Relevant Variants in SARS-CoV-2 and *Mycobacterium tuberculosis* Co-infection": https://docs.google.com/document/d/1Qg9nVQFBlbtU1rxPrbDtTWPhalmwwtRB/edit?usp=sharing&ouid=108297540105191031878&rtpof=true&sd=true

Supplementary Tables

Supplementary Table 1: Single nucleotide polymorphisms (SNPs) in the ACE2 gene showing significant associations (P < 0.05) with IL-1β, IL-6, IFN-γ, and TNF-α

| SNP | Cytokine | P-Value |
| --- | --- | --- |
| Rs6632677 | Il-1β | 0.0454 |
| Rs147311723 | IL-6 | 0.0471 |
| Rs4646140 | IL-6 | 0.0364 |
| Rs2285666 | IFN-γ | 0.0465 |
| Rs2285666 | TNF-α | 0.0194 |
| Rs4646142 | TNF-α | 0.0472 |
| Rs6632677 | TNF-α | 0.0091 |

Supplementary Table 2: Allele distribution of single nucleotide polymorphisms (SNPs) of ACE2 showing significant associations with biochemical markers.

| SNP | Variable | P-Value |
| --- | --- | --- |
| rs147311723 | Crea | 0.0522 |
| rs2285666 | AST | 0.0536 |
| rs4646142 | ALT | 0.0582 |
